# Challenges to self-isolation among contacts of cases of COVID-19: a national telephone survey in Wales

**DOI:** 10.1101/2021.07.23.21261020

**Authors:** Kate R Isherwood, Richard G Kyle, Benjamin J Gray, Alisha R Davies

**Affiliations:** Research and Evaluation Division, Public Health Wales, Capital Quarter 2, Cardiff, Wales, CF10 4BZ

**Keywords:** COVID-19, self-isolation, challenges, health inequalities, financial stability

## Abstract

**Objectives:** To identify the specific challenges of self-isolation experienced by population sub-groups to better target and tailor support.

**Design:** The **C**ontact **A**dherence **B**ehavioural **In**sights **S**tudy (CABINS) was a 15-minute telephone survey of confirmed contacts of cases of COVID-19 identified through the national NHS Wales Test Trace Protect (TTP) database.

**Methods:** Confirmed contacts of cases of COVID-19 reached by TTP completed a 15-minute telephone survey (N = 2,027). Binary logistic regression models adjusted for age, gender, living alone, survey round, deprivation quintile (defined by the Welsh Index of Multiple Deprivation) and income precarity (financial security) determined which population sub-groups were more likely to experience challenges during self-isolation.

**Results:** Younger people (aged 18-29 years) were 3 times more likely to report mental health concerns (Adjusted Odds Ratio [aOR]: 3.16, 95% Confidence Interval [CI]: 2.05-4.86) and 2 times more likely to report loneliness (aOR: 1.96, CI: 1.37-2.81) compared to people aged over 60 years. Women were 1.5 times more likely to experience mental health concerns (aOR: 1.51, 95% CI: 1.20-1.92) compared to men. People with high/very high levels of income precarity were 8 times more likely to report financial challenges (aOR: 7.73, CI: 5.10-11.74) and 3 times more likely to report mental health concerns than their more financially secure counterparts (aOR: 3.08, CI: 2.22-4.28).

**Conclusions:** Self-isolation is particularly challenging for those with younger people, women and precarious incomes. Providing enhanced emotional, financial and social support and signposting to these groups is required to minimise the harms of self-isolation.

**What is already known on this subject?:**

- Self-isolation after notification as a contact of a positive case of COVID-19 is essential to prevent the spread of the disease. However, self-isolation can be challenging and adherence is dependent upon a range of psychological, social and economic factors.
- Emerging data suggests that the COVID-19 pandemic is having disproportionate impact on those on lower incomes and those of lower socio-economic status.

**What does this study add?:**

- The most common challenges faced to self-isolation were wanting to see family and friends, followed by a lack of exercise.
- Individuals with some income precarity were more than 7 times more likely to report financial concerns and 3 times more likely to report mental health concerns as a challenge to self-isolation than those who were financially secure.
- Interventions to support individuals to self-isolate needs to be targeted at groups most susceptible to experiencing challenges to self-isolation for infectious diseases. Our research suggests that the use of income precarity questions as a screening tool is important to direct financial and practical support through contact tracing systems.

## INTRODUCTION

Self-isolation of close contacts of confirmed cases of COVID-19 has been a key measure to stop community spread of Coronavirus (SARS-CoV-2) across a number of countries^[1]^. In the United Kingdom (UK), self-isolation has been a legal requirement for confirmed contacts since 28^th^ September, 2020^[2]^. Self-isolation for a period of 14 days was required until 9^th^ December 2020, after which the duration of self-isolation reduced to 10 days^[3]^. Although differences in service delivery exist across the UK ^[4-7]^, in each UK nation contact tracing of index cases is used to identify their close contacts who are then informed to self-isolate. In Wales, contact tracing is conducted through NHS Wales Test, Trace, Protect (TTP) by local contact tracing teams working in each of the 22 Local Authorities across Wales^[5]^. TTP is supported by a national database that records details of all cases and their identified contacts who are successfully traced and informed to self-isolate^[5]^. For the period since contact tracing in Wales by TTP started on 21^st^ June 2020 up to 17^th^ July, 2021, 439,753 close contacts of cases have been identified, of which 417,703 (95.0%) have been successfully contacted and informed to self-isolate^[8]^.

Studies report adherence to self-isolation between 11% and 86% in the UK ^[9-11]^. However, despite this variation in the levels of self-reported adherence, roughly 80% report experiencing challenges to adhering to the UK Government’s COVID-19 related instructions^[12]^. The most prevalent challenges reported were mental health concerns, physical health difficulties and changes in daily routine^[12]^. Men and those aged 55 years or older are least likely to report challenges^[12]^.

Identifying and understanding the challenges faced during self-isolation is important to help inform, target and tailor the support needed both during the COVID-19 pandemic and to minimise harms of future infectious disease outbreaks. Nevertheless, evidence on the challenges experienced during self-isolation among contacts of cases of COVID-19 is limited. As a single national digital platform exists in Wales to support contact tracers, this provides an opportunity to provide robust evidence for the rest of the UK and internationally. Our study aimed to identify the specific challenges experienced by population sub-groups to better develop and target the support needed to minimise potential health and economic harms of self-isolation.

## METHODS

### Study Design

The COVID-19 **<u>C</u>**ontacts **<u>A</u>**dherence **<u>B</u>**ehavioural **<u>In</u>**sights **<u>S</u>**urvey (CABINS) was a mixed-methods study of confirmed contacts of cases of COVID-19 reached by TTP. CABINS included two rounds of a telephone survey followed by online focus groups with a sample of telephone survey participants after each round. Round 1 was undertaken between 11^th^ November and 1^st^ December, 2020 and Round 2 between 18^th^ February and 23^rd^ March, 2021. Here we report telephone survey data from both rounds of data collection.

### Participants

Individuals were eligible for inclusion during each survey round if they: (1) had been successfully contacted by TTP after forward contact tracing^1^ and informed to self-isolate; (2) were a close contact of a confirmed case of COVID-19; (3) were aged 18 years or over; (4) resident in Wales; and (5) had completed their self-isolation period at the time of telephone survey. Contacts were excluded from the study if they were: (1) under the age of 18; (2) currently self-isolating; (3) not a resident in Wales; or (4) a contact of a case of COVID-19 who died.

We used the national TTP database to identify eligible participants who have been asked to self-isolate at two different time periods (*n =* 47,072). Round 1 was between 12^th^ September 2020 and 23^rd^ October 2020, the day before the start of the Welsh ‘Firebreak’^[13]^ (*n* = 18,568). Round 2 was between 13^th^ December and 15^th^ January 2021 (*n* = 28,504). Due to changes in Government guidelines^[14]^ the legally required duration of self-isolation during Round 1 was 14 days, and during Round 2 was 10 days.

#### Sampling

A target sample size of 1,000 was set for each survey round. This threshold was set to enable a sufficient number of participants to enable sub-group analysis. Quota sampling was applied based on age and gender (combined) and Welsh Index of Multiple Deprivation (WIMD) quintile to ensure that the sample was representative of contacts informed to self-isolate during each period.

#### Recruitment

Figure 1 shows the study flowchart. Eligible participants were invited to participate in a 15-minute telephone interview. Bilingual interviews were conducted in either Welsh or English according to participant preference. We used an external market research company (Beaufort Research Limited) to recruit participants and collect data to ensure anonymity and minimise potential response bias if Public Health Wales had made initial contact with participants. Participants were approached for interview between 12^th^ November and 1^st^ December, 2020 for those asked to self-isolate during Round 1 and between 18^th^ February and 13^th^ March, 2021 for Round 2. Each respondent was contacted up to a maximum of seven times. Across both survey rounds, 47,072 contacts were eligible to participate. 10,801 (23.0%) were approached by telephonist and 2,027 (18.8%) completed the telephone survey.

**Figure 1:**
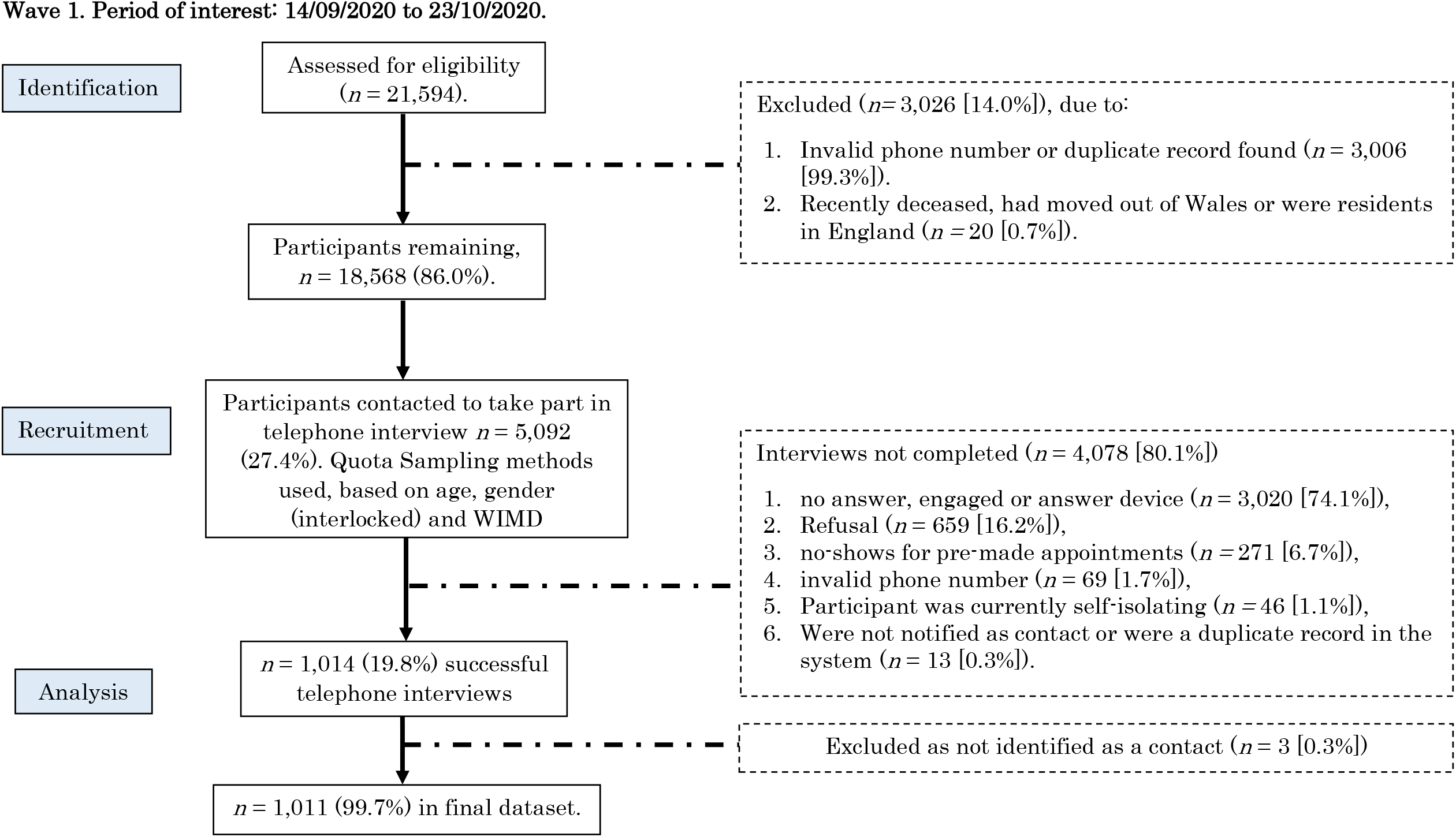
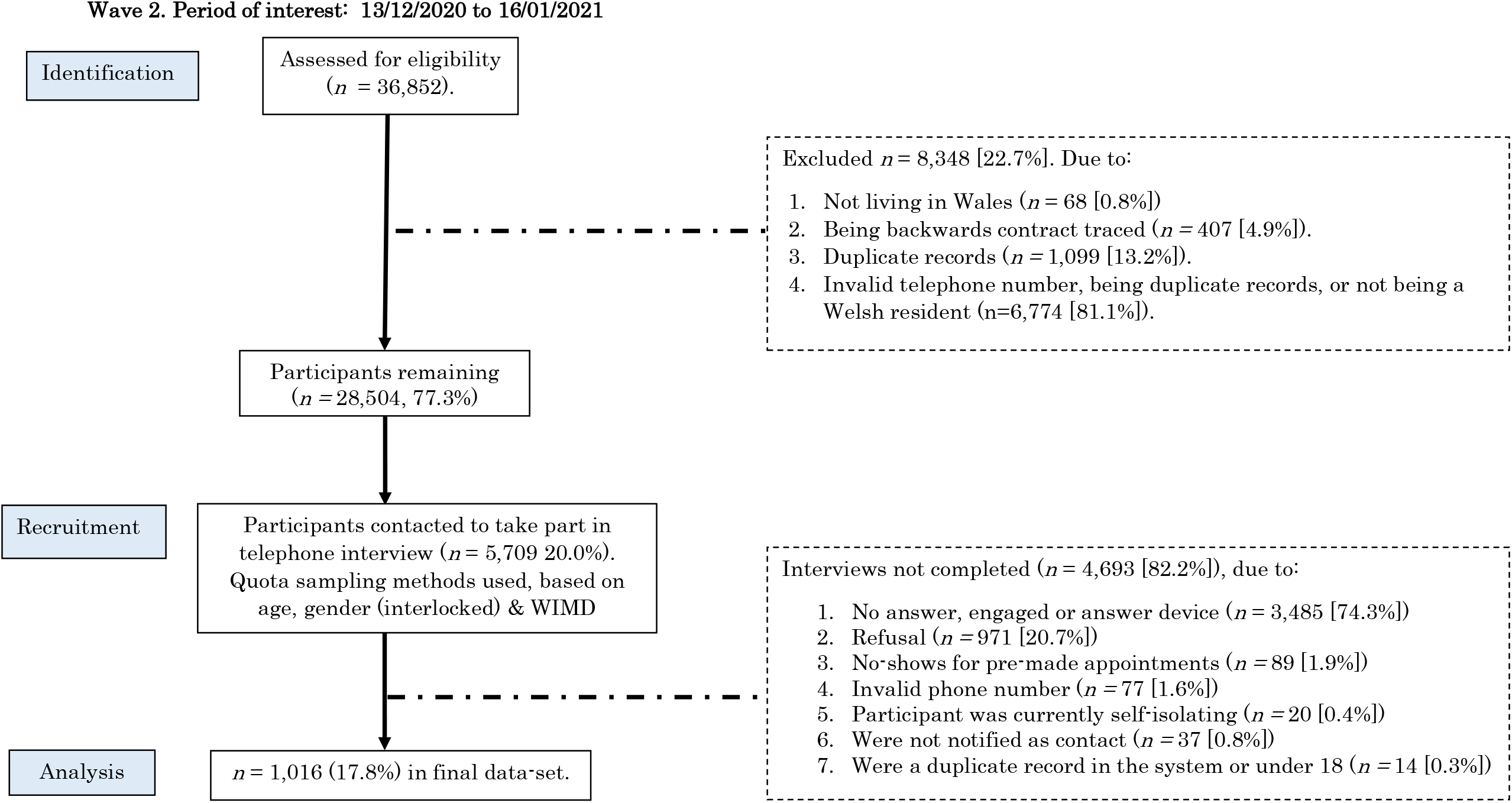
Study flowchart for both waves.

### Measures

#### Challenges to self-isolation

Participants were asked to select from a list of challenges experienced during self-isolation. In Round 1 this list included 15 options. Five additional options were added to Round 2, derived from data analysis on this question conducted after Round 1 and from the companion real-time Adherence Confidence Text Survey (ACTS)^[10]^. Challenges were treated as a binary ‘selected’ or ‘not selected’ variable in analysis.

#### Income precarity

To assess income precarity, questions were taken from the Wages subscale of the Employment Precariousness Scale (EPRES)^[14]^. Participants were asked to provide their income during their self-isolation period using one of five categories (ranging from ‘Less than £200 a year/ less than £870 a month/ less than £10,400 a year’ to ‘£800 or more a week/ £3,460 or more a month/ £41,500 or more a year’), whether this income enabled them to cover their basic needs, and to what extent it enabled them to cover unforeseen expenses, on a 5-point Likert Scale (from ‘always’ to ‘never’). A ‘don’t know’ and ‘prefer not to say’ option was added to all questions for those who did not want to disclose their financial situation. This 5-point Likert Scale was then recoded onto a 0-4 scale (where 0 = Always, 4 = Never). Scores for each item on the scale was divided by 12, summed and multiplied by 4 to give a composite Precarious Income (Wages) score^[14]^. A composite score below 1 indicates low income precarity (i.e., are more financially secure); between 1 and 1.99 is moderate income precarity; and a score of 2 and above is considered high and very high precarious income precarity^[14]^.

#### Socio-demographic characteristics

Socio-demographic characteristics collected included age, gender, ethnicity and living alone. Participant’s WIMD quintile was derived using postcode data held by TTP.

### Data analysis

Analysis was conducted in three steps. First, descriptive statistics were calculated for all variables and reported as n(%). Second, chi-squared tests were conducted to assess associations between socio-demographic characteristics and survey round, and between reported challenges to self-isolation and sociodemographic characteristics. Third, binary logistic regression models, adjusted for age, gender, WIMD, living alone, survey round and income precarity were built to assess independent predictors of each challenge reported as Adjusted Odds Ratios (aOR) with 95% Confidence Intervals (CI).

Weights were applied to all analyses to ensure that the final sample was representative of all eligible participants in each survey round. A sensitivity analysis was conducted using the data for each survey round separately and showed no difference in results. To increase statistical power, analysis is therefore reported on the full study sample (n=2,027), adjusting for survey round. Due to the number of logistic regression models built, Bonferroni correction was applied to account for multiple testing and significance set at p≤0.002 (i.e., p<0.05/20 reported challenges). Only those findings that are significant at the corrected level are reported in the results narrative.

### Ethical considerations

The Health Research Authority approved the CABINS study (IRAS: 289377).

## RESULTS

### Sample characteristics

Table 1 displays sample characteristics. There were more female participants (53.6%) than male, over two-fifths lived in the most deprived quintiles (45.1%) and three-fifths (62.9%) had some degree of income precarity (i.e., moderate or high/very high).

**Table 1.** Telephone Survey Sample Characteristics by survey round

|  | All respondents |  | Survey Round |  |  |  | Sig. |
| --- | --- | --- | --- | --- | --- | --- | --- |
|  |  |  | 1 |  | 2 |  |  |
|  | N = 2,027 |  | N= 1,011 |  | N= 1,016 |  |  |
|  | % | N | % | N | % | N |  |
| **Gender** <sup>[1]</sup> |  |  |  |  |  |  |  |
| Male | 45.9 | 931 | 46.9 | 472 | 45.4 | 459 | .50 |
| Female | 53.6 | 1086 | 53.1 | 533 | 54.6 | 553 |  |
| **Age** <sup>[2]</sup> |  |  |  |  |  |  |  |
| 18-29 | 29.5 | 598 | 31.8 | 322 | 26.9 | 273 | .13 |
| 30-39 | 18.5 | 374 | 17.7 | 179 | 18.8 | 190 |  |
| 40-49 | 17.0 | 344 | 16.1 | 163 | 18.1 | 183 |  |
| 50-59 | 20.0 | 405 | 19.5 | 197 | 21.0 | 213 |  |
| 60+ | 9.6 | 301 | 8.9 | 150 | 10.4 | 155 |  |
| **WIMD** <sup>[3]</sup> |  |  |  |  |  |  |  |
| 1 – Most Deprived | 21.6 | 437 | 22.1 | 223 | 21.1 | 214 | .41 |
| 2 | 23.5 | 476 | 22.7 | 230 | 24.1 | 245 |  |
| 3 | 20.4 | 413 | 19.4 | 196 | 21.7 | 220 |  |
| 4 | 17.7 | 359 | 17.5 | 177 | 17.5 | 178 |  |
| 5 – Least Deprived | 16.9 | 342 | 18.3 | 185 | 15.6 | 159 |  |
| **Income Precarity** <sup>[5]</sup> |  |  |  |  |  |  |  |
| Low | 22.7 | 460 | 26.3 | 229 | 26.7 | 231 | .72 |
| Moderate | 34.5 | 700 | 39.7 | 344 | 41.1 | 356 |  |
| High/ Very High | 28.4 | 575 | 34.0 | 295 | 32.2 | 279 |  |
| **Living alone** <sup>[6]</sup> |  |  |  |  |  |  |  |
| Yes | 18.4 | 372 | 20.1 | 203 | 16.7 | 169 | .05 |
| No | 81.6 | 1655 | 79.9 | 808 | 83.3 | 847 |  |

29. **Notes:** [1] 10 people excluded from analysis who chose 'In another way' or 'Prefer not to say'; [2] 2 people excluded from analysis who failed to provide their age; [3] Welsh Index of Multiple Deprivation; [4] Defined as those who self-reported that they were either: (1) a student, (2) long-term sick or disabled, (3) retired, or (4) a carer; [5] 293 excluded from analysis due to incomplete data; [6] defined as living with no other adults. Figures in bold represent statistical significance (to p < .05).

### Challenges reported to self-isolation

#### Challenges faced to self-isolation

‘Wanting to see family’ (Round 1 = 66.7%; Round 2= 61.9% [Table 2]) and ‘wanting to see friends’ (Round 1 = 60.6%; Round 2 = 58.2% [Table 2]) were the main challenges to self-isolation in both survey rounds. Around a third of individuals reported that loneliness was a challenge to self-isolation (Round 1 = 31.2% vs. Round 2 = 27.3% [Table 2]) and a quarter of the sample in each survey round said that mental health difficulties were a challenge to self-isolation (Round 1 = 24.6% vs. Round 2 = 24.8% [Table 2]).

**Table 2:** Challenges faced to self-isolation, split by survey round.

| Challenge | All respondents |  | Survey Round |  |  |  | Sig |
| --- | --- | --- | --- | --- | --- | --- | --- |
|  |  |  | 1 |  | 2 |  |  |
|  | N = 2,027 |  | N= 1,011 |  | N=1,016 |  |  |
|  | % | N | % | N | % | N |  |
| You wanted to see family | 64.3 | 1303 | 66.7 | 674 | 61.9 | 629 | .03 |
| You wanted to see friends | 59.4 | 1205 | 60.6 | 613 | 58.2 | 592 | .28 |
| A lack of exercise/ fresh air | 51.0 | 1033 | 58.6 | 592 | 43.4 | 441 | <.001 |
| Loneliness | 29.3 | 593 | 31.2 | 315 | 27.3 | 278 | .06 |
| Mental health challenges | 24.7 | 501 | 24.6 | 249 | 24.8 | 252 | .93 |
| Financial concerns | 18.5 | 376 | 20.4 | 206 | 16.7 | 170 | .04 |
| Caring responsibilities for vulnerable adults outside your household | 17.2 | 348 | 17.7 | 173 | 17.3 | 175 | .95 |
| Living with people who are not self-isolating | 14.7 | 297 | 17.5 | 177 | 11.8 | 120 | <.001 |
| No access to food | 9.9 | 201 | 7.7 | 78 | 12.1 | 123 | .001 |
| Caring for an individual within your household <sup>[1]</sup> | 10.1 | 204 | - | - | 20.1 | 204 | - |
| Work not supporting you to self-isolate | 7.3 | 148 | 8.8 | 89 | 5.8 | 59 | .01 |
| Physical health challenges | 9.8 | 198 | 9.5 | 96 | 10.0 | 102 | .68 |
| Caring responsibilities for children outside your household | 8.4 | 171 | 8.5 | 86 | 8.4 | 85 | .91 |
| Concerns about the impact of isolation on work or business <sup>[1]</sup> | 7.9 | 160 | - | - | 15.8 | 160 | - |
| Looking after pets or animals <sup>[1]</sup> | 8.8 | 179 | - | - | 17.6 | 179 | - |
| Self-isolating away from vulnerable household members <sup>[1]</sup> | 8.3 | 168 | - | - | 16.5 | 168 | - |
| Lack of support from family and friends | 5.8 | 119 | 64 | 63 | 5.5 | 56 | .44 |
| A lack of private outdoors space <sup>[1]</sup> | 5.2 | 105 | - | - | 10.3 | 105 | - |
| No access to medication | 4.8 | 98 | 5.3 | 54 | 4.4 | 44 | .29 |
| You did not feel safe to isolate | 1.3 | 26 | 1.6 | 16 | 1.0 | 10 | .23 |
| None of the above | 10.2 | 206 | 9.6 | 97 | 10.7 | 109 | .40 |

Notes: [1] options only available in Wave 2 Telephone Survey. Figures in bold represent statistical significance (corrected to p = .002).

#### Differences between survey rounds

Individuals were significantly less likely to say ‘a lack of exercise’ was a challenge in Round 2 than Round 1 (Round 2 = 43.4% vs Round 1 = 58.6, p = <.001; [Table 2]). A significantly greater number of individuals reported that having ‘no access to food’ was a challenge to self-isolation in Round 2 than Round 1 (Round 2 = 12.1% vs Round 1 = 7.7%; p = .001; [Table 2]). Individuals were also significantly more likely to say that ‘living with others who were not self-isolating’ was a challenge in Round 1 than in Round 2 (Round 2 = 11.8% vs. Round 1 = 17.5%, p = <.001; [Table 2]).

### Challenges to self-isolation by socio-demographic group

#### Age

In logistic regression models, adjusted for age, gender, living alone, survey round, and WIMD, young people (aged 18-29) were 3 times more likely to report a lack of exercise (aOR = 3.51, [95% CI =2.50-4.93]), mental health difficulties (aOR = 3.16, [95% CI =2.05-4.86]) and concerns about the impact of self-isolation on work or business (aOR = 3.46, [95% CI =1.63-7.35]) than older people [Table 3]. Young people (aged 18-29) were also 2 times more likely to report wanting to see friends (aOR = 2.25, [95% CI =1.62-3.21]) and loneliness (aOR = 1.96, [95% CI =1.37-2.81]) as challenges to self-isolation, compared to older people. Those under the age of 39 years were 3 times more likely to report mental health difficulties (18-29: aOR = 3.16, [95% CI =2.05-4.86]; 30-39: aOR = 2.46, [95% CI =1.55-3.91]) and those aged between 30 and 49 years were 3 times more likely to report financial concerns as a challenge to self-isolation (30-39: aOR = 2.90, [95% CI =1.72-4.89; 40-49: (aOR = 2.78, [95% CI =1.62-4.76] [Table 3]). Individuals aged 30 to 39 years were 3 times more likely to have caring responsibilities for a vulnerable individual in the household (aOR = 3.32, [95% CI =1.71-6.45]), whereas those aged 50 to 59 years where 2 times more likely to say they had caring responsibilities for vulnerable adults outside the household (aOR = 2.36, [95% CI =1.50-3.74]) [Table 3].

**Table 3:** Logistic Regression Models

| Rank | Challenge | N | Age |  |  |  |  | Gender |  | Living alone |  | Survey Round |  |
| --- | --- | --- | --- | --- | --- | --- | --- | --- | --- | --- | --- | --- | --- |
|  |  |  | 18-29 | 30-39 | 40-49 | 50-59 | 60+ | Male | Female | Yes | No | 1 | 2 |
| 1 | Wanting to see family | 1136 | 1.07<br>[.76-1.51] | .85<br>[.59-1.22] | .66<br>[.46-.96] | .76<br>[.53-1.09] | Ref. | Ref. | 1.35<br>[1.10-1.66] | .91<br>[.70-1.20] | Ref. | Ref. | .86<br>[.71-1.06] |
| 2 | Wanting to see friends | 995 | **2.25**<br>[**1.62-3.12**] | 1.20<br>[.85-1.69] | 1.07<br>[.75-1.51] | 1.01<br>[.82-1.23] | Ref. | Ref. | 1.01<br>[.82-1.23] | .81<br>[.62-1.05] | Ref. | Ref. | .96<br>[.79-1.17] |
| 3 | A lack of exercise | 287 | **3.51**<br>[**2.50-4.93**] | **3.01**<br>[**2.10-4.32**] | **2.08**<br>[**1.44-2.99**] | 1.59<br>[1.11-2.26] | Ref. | Ref. | 1.02<br>[.83-1.25] | .81<br>[.62-1.09] | Ref. | Ref. | **.52**<br>[**.42-.63**] |
| 4 | Loneliness | 507 | **1.96**<br>[**1.37-2.81**] | 1.18<br>[.80-1.75] | .98<br>[.65-1.48] | .91<br>[.61-1.36] | Ref. | Ref. | 1.31<br>[1.05-1.64] | **.41**<br>[**.31-.53**] | Ref. | Ref. | .87<br>[.70-1.08] |
| 5 | Mental health difficulties | 374 | **3.16**<br>[**2.05-4.86**] | **2.46**<br>[**1.55-3.91**] | 1.95<br>[1.21-3.15] | 1.61<br>[1.00-2.59] | Ref. | Ref. | **1.51**<br>[**1.20-1.92**] | .71<br>[.53-.96] | Ref. | Ref. | 1.04<br>[.82-1.30] |
| 6 | Financial concerns | 327 | 2.10<br>[1.28-3.45] | **2.90**<br>[**1.72-4.89**] | **2.78**<br>[**1.62-4.76**] | 2.08<br>[1.22-3.55] | Ref. | Ref. | .71<br>[.54-.92] | .91<br>[.65-1.27] | Ref. | Ref. | .84<br>[.66-1.09] |
| 7 | Caring responsibilities for vulnerable adults outside your household | 304 | 1.05<br>[.66-1.67] | 1.21<br>[.74-1.98] | 1.87<br>[1.59-3.00] | **2.36**<br>[**1.50-3.74**] | Ref. | Ref. | 1.27<br>[.98-1.65] | 1.03<br>[.80-1.32] | Ref. | Ref. | 1.03<br>[.80-1.32] |
| 8 | Living with others who are not self-isolating | 266 | 2.02<br>[1.19-3.45] | 2.19<br>[1.25-3.85] | 2.29<br>[1.30-4.03] | 1.69<br>[.96-2.99] | Ref. | Ref. | 1.12<br>[.85-1.47] | **2.71**<br>[**1.70-4.33**] | Ref. | Ref. | **.62**<br>[**.48-.82**] |
| 9 | No access to food | 181 | 1.51<br>[.87-2.63] | 1.39<br>[.77-2.52] | 1.38<br>[.75-2.53] | 1.00<br>[.54-1.86] | Ref. | Ref. | .98<br>[.71-1.35] | 1.66<br>[.96-2.88] | Ref. | Ref. | 1.57<br>[1.14-2.16] |
| 10 | Caring for a vulnerable individual in the household [1] | 170 | 1.21<br>[.62-2.39] | **3.32**<br>[**1.71-6.45**] | 1.95<br>[.97-3.92] | 1.45<br>[.72-2.91] | Ref. | Ref. | .93<br>[.65-1.32] | 1.60<br>[.95-2.69] | Ref. | Ref. | - |
| 12 | Physical health difficulties | 163 | .73<br>[.43-1.25] | .74<br>[.41-1.34] | 1.21<br>[.69-2.12] | 1.05<br>[.61-1.84] | Ref. | Ref. | .74<br>[.53-1.04] | .70<br>[.47-1.05] | Ref. | Ref. | 1.04<br>[.75-1.44] |
| 13 | Looking after pets [1] | 159 | 3.14<br>[1.53-6.48] | 1.67<br>[.76-3.65] | 2.25<br>[1.04-4.88] | 2.59<br>[1.23-5.48] | Ref. | Ref. | 1.36<br>[.95-1.96] | 1.66<br>[.96-2.88] | Ref. | Ref. | - |
| 14 | Caring responsibilities for children outside your household | 155 | **.34**<br>[**.19-.62**] | .77<br>[.44-1.38] | 1.01<br>[.57-1.78] | 1.28<br>[.76-2.17] | Ref. | Ref. | .91<br>[.65-1.28] | 1.30<br>[.82-2.05] | Ref. | Ref. | 1.57<br>[1.14-2.16] |
| 15 | Self-isolating away from vulnerable household members | 148 | 1.55<br>[.78-3.06] | 1.84<br>[.92-3.72] | 1.63<br>[.80-3.36] | 1.44<br>[.71-2.92] | Ref. | Ref. | 1.10<br>[.76-1.59] | 1.80<br>[1.02-3.19] | Ref. | Ref. | - |
| 16 | Concerned about impact of isolation on work or business [1] | 141 | **3.46**<br>[**1.63-7.35**] | 2.02<br>[.90-4.54] | 2.38<br>[1.06-5.38] | 1.55<br>[.68-3.55] | Ref. | Ref. | .56<br>[.38-.82] | .97<br>[.57-1.65] | Ref. | Ref. | - |
| 17 | Work not supporting you to self-isolate | 139 | 1.93<br>[.98-3.83] | 1.96<br>[.95-4.04] | 1.55<br>[.72-3.35] | 1.75<br>[.83-3.66] | Ref. | Ref. | .60<br>[.42-.87] | .72<br>[.46-1.12] | Ref. | Ref. | .70<br>[.49-1.00] |
| 18 | Lack of support from family/ friends | 104 | 1.42<br>[.66-3.04] | 2.25<br>[1.05-4.83] | 1.92<br>[.86-4.26] | 1.42<br>[.63-3.19] | Ref. | Ref. | .64<br>[.43-.98] | **.38**<br>[**.24-.59**] | Ref. | Ref. | .84<br>[.56-1.26] |
| 19 | A lack of private outdoors space [1] | 91 | 3.09<br>[1.36-7.02] | 1.20<br>[.47-3.04] | 1.40<br>[.55-3.54] | 1.40<br>[.55-3.54] | Ref. | Ref. | .61<br>[.39-.97] | .72<br>[.39-1.31] | Ref. | Ref. | - |
| 20 | No access to medication | 86 | 1.11<br>[.52-2.36] | 1.07<br>[.47-2.45] | 1.23<br>[.54-2.80] | 1.08<br>[.47-2.45] | Ref. | Ref. | 1.14<br>[.73-1.79] | 1.31<br>[.71-2.44] | Ref. | Ref. | .84<br>[.54-1.31] |
| 21 | Not feeling safe to self-isolate | 23 | .77<br>[.20-2.98] | 1.17<br>[.28-4.92] | .54<br>[.09-3.28] | .93<br>[.21-4.22] | Ref. | Ref. | .49<br>[.21-1.16] | 1.45<br>[.42-5.03] | Ref. | Ref. | .55<br>[.23-1.31] |
| 11 | None of the above | 164 | **.36**<br>[**.22-.59**] | **.38**<br>[**.22-.66**] | .60<br>[.36-1.00] | .45<br>[.27-.75] | Ref. | Ref. | .90<br>[.65-1.26] | 1.41<br>[.89-2.23] | Ref. | Ref. | 1.24<br>[.90-1.73] |

Notes: [1] options only available in Wave 2 Telephone Survey. Figures in bold represent statistical significance (corrected to p =.002).

**Table 3 continued:** Logistic Regression Table
| Challenge | N | WIMD |  |  |  |  | Income Precarity<sup>[2]</sup> |  |  |
| --- | --- | --- | --- | --- | --- | --- | --- | --- | --- |
|  |  | 1 | 2 | 3 | 4 | 5 | Low | Moderate | High/ V. High |
| 1 Wanting to see family | 1136 | .93<br>[.67-1.29] | 1.09<br>[.79-1.51] | 1.12<br>[.81-1.56] | 1.32<br>[.93-1.87] | Ref. | Ref. | 1.02<br>[.79-1.32] | 1.12<br>[.85-1.47] |
| 2 Wanting to see friends | 995 | 1.02<br>[.74-1.42] | 1.01<br>[.74-1.38] | .94<br>[.68-1.29] | 1.18<br>[.84-1.65] | Ref. | Ref. | .96<br>[.74-1.23] | 1.24<br>[.95-1.62] |
| 3 A lack of exercise | 287 | .69<br>[.50-.97] | .60<br>[.43-.83] | .70<br>[.50-.97] | .59<br>[.42-.82] | Ref. | Ref. | .84<br>[.65-1.09] | 1.20<br>[.92-1.57] |
| 4 Loneliness | 507 | .84<br>[.59-1.20] | 1.07<br>[.76-1.52] | .95<br>[.66-1.35] | .81<br>[.56-1.18] | Ref. | Ref. | 1.30<br>[.97-1.75] | **2.11**<br>**[1.56-2.85]** |
| 5 Mental health difficulties | 374 | 1.20<br>[.82-1.76] | 1.24<br>[.85-1.80] | 1.20<br>[.82-1.77] | .79<br>[.52-1.20] | Ref. | Ref. | 1.40<br>[1.00-1.95] | **3.08**<br>**[2.22-4.28]** |
| 6 Financial concerns | 327 | 1.79<br>[1.15-2.78] | 1.62<br>[1.04-2.51] | 1.43<br>[.91-2.26] | 1.18<br>[.73-1.90] | Ref. | Ref. | **2.56**<br>**[1.67-3.92]** | **7.73**<br>**[5.10-11.74]** |
| 7 Caring responsibilities for vulnerable adults outside your household | 304 | 1.19<br>[.78-1.82] | 1.27<br>[.85-1.91] | 1.19<br>[.79-1.81] | .97<br>[.62-1.52] | Ref. | Ref. | .82<br>[.60-1.14] | 1.07<br>[.77-1.50] |
| 8 Living with others who are not self-isolating | 266 | 1.05<br>[.67-1.63] | 1.19<br>[.77-1.82] | .96<br>[.61-1.50] | 1.08<br>[.69-1.69] | Ref. | Ref. | .90<br>[.64-1.28] | 1.04<br>[.73-1.49] |
| 9 No access to food | 181 | .98<br>[.59-1.62] | .77<br>[.46-1.30] | 1.09<br>[.66-1.80] | .78<br>[.45-1.35] | Ref. | Ref. | .89<br>[.57-1.38] | 1.90<br>[1.25-2.89] |
| 10 Caring for a vulnerable individual in the household <sup>[1]</sup> | 170 | 1.18<br>[.67-2.10] | 1.09<br>[.62-1.93] | .99<br>[.56-1.77] | .74<br>[.39-1.41] | Ref. | Ref. | 1.16<br>[.72-1.86] | **2.51**<br>**[1.57-4.03]** |
| 12 Physical health difficulties | 163 | 1.18<br>[.70-1.98] | .84<br>[.49-1.44] | .93<br>[.55-1.60] | .67<br>[.37-1.23] | Ref. | Ref. | 1.94<br>[1.19-3.14] | **2.57**<br>**[1.57-4.21]** |
| 13 Looking after pets <sup>[1]</sup> | 159 | 1.30<br>[.74-2.31] | .77<br>[.43-1.40] | 1.06<br>[.60-1.89] | .86<br>[.46-1.59] | Ref. | Ref. | 1.14<br>[.72-1.82] | 1.30<br>[.80-2.11] |
| 14 Caring responsibilities for children outside your household | 155 | 1.18<br>[.68-2.04] | 1.17<br>[.69-1.99] | .77<br>[.43-1.38] | .96<br>[.53-1.72] | Ref. | Ref. | 1.52<br>[.94-2.43] | **2.39**<br>**[1.48-3.84]** |
| 15 Self-isolating away from vulnerable household members | 148 | .83<br>[.46-1.49] | .69<br>[.38-1.23] | 1.00<br>[.57-1.76] | .64<br>[.34-1.20] | Ref. | Ref. | .94<br>[.59-1.50] | 1.47<br>[.91-2.36] |
| 16 Concerned about impact of isolation on work or business <sup>[1]</sup> | 141 | .98<br>[.51-1.86] | 1.30<br>[.70-2.41] | .89<br>[.46-1.70] | 1.29<br>[.67-2.49] | Ref. | Ref. | 1.25<br>[.75-2.10] | 2.26<br>[1.35-3.78] |
| 17 Work not supporting you to self-isolate | 139 | 2.10<br>[1.13-3.89] | 1.51<br>[.80-2.85] | 1.35<br>[.70-2.61] | 1.09<br>[.54-2.21] | Ref. | Ref. | 1.31<br>[.76-2.26] | **2.99**<br>**[1.77-5.02]** |
| 18 Lack of support from family/ friends | 104 | .82<br>[.44-1.55] | .85<br>[.46-1.58] | .82<br>[.43-1.57] | .38<br>[.16-.87] | Ref. | Ref. | .98<br>[.54-1.77] | **2.45**<br>**[1.41-4.25]** |
| 19 A lack of private outdoors space <sup>[1]</sup> | 91 | 1.84<br>[.83-4.10] | 1.62<br>[.72-3.61] | 1.16<br>[.50-2.69] | 1.44<br>[.61-3.40] | Ref. | Ref. | 1.17<br>[.62-2.23] | 1.95<br>[1.04-3.66] |
| 20 No access to medication | 86 | 1.29<br>[.66-2.52] | .79<br>[.39-1.63] | 1.10<br>[.55-2.20] | .39<br>[.16-.99] | Ref. | Ref. | .92<br>[.49-1.74] | 1.88<br>[1.03-3.43] |
| 21 Not feeling safe to self-isolate | 23 | 2.17<br>[.45-10.50] | 1.19<br>[.21-6.58] | 2.27<br>[.46-11.33] | 1.13<br>[.19-6.86] | Ref. | Ref. | 4.86<br>[.60-39.44] | 12.68<br>[1.66-97.00] |
| 11 None of the above | 164 | 1.07<br>[.61-1.87] | 1.44<br>[.85-2.43] | 1.35<br>[.80-2.29] | .86<br>[.47-1.57] | Ref. | Ref. | .74<br>[.50-1.09] | .55<br>[.35-.87] |

Notes: [1] options only available in Wave 2 Telephone Survey. Figures in bold represent statistical significance (corrected to p = .002). ; [2] Not all respondents answered the 3 questions associated with the income precarity measure (N = 292), leaving a sample of 1735.

#### Gender

Women were also 1.5 times more likely than men to say they experienced mental health difficulties as a challenge to self-isolation (aOR = 1.51, [95% CI =1.20-1.92]) [Table 3].

#### Income precarity

Individuals with moderate income precarity were 2 times more likely to report financial concerns as a challenge to self-isolation (aOR = 2.56 [95% CI = 1.67-3.92]) than their more financially secure counterparts [Table 3]. Those who were in the highest income precarity group were over 7 times more likely to report financial concerns (aOR = 7.73 [95% CI = 5.10-11.74]) and were 3 times more likely to report mental health difficulties (aOR = 3.08 [95% CI = 2.22-4.28]), caring for a vulnerable individual in the household (aOR = 2.51, [95% CI = 1.57-4.03]), physical health difficulties (aOR = 2.57, [95% CI =1.57-4.21]) and work not supporting them to self-isolate (aOR = 2.99, [95% CI = 1.77-5.02]) as a challenge to self-isolation [Table 3]. They were also 2 times more likely to report a lack of support from family or friends (aOR = 2.45, [95% CI = 1.41-4.25]), caring responsibilities for children outside the household (aOR = 2.39, [95% CI = 1.48-3.84]) and loneliness (aOR = 2.11, [95% CI = 1.56-2.85]) [Table 3]. No significant differences were found when looking at the differences in the challenges reported by deprivation quintile [Table 3].

## DISCUSSION

Our study identified that self-isolation had a disproportionate impact on different population groups, particularly, people of younger age (aged 18-29), women and those with precarious incomes.

### Challenges faced to self-isolation by young people

Our findings suggest that self-isolation is particularly challenging for younger people aged under 29. This population were more likely to report mental health difficulties, loneliness, and concerns about the impact of self-isolation on their work or business, wanting to see friends and a lack of exercise as challenges to self-isolation.

Young people are suffering serious psychosocial effects of the COVID-19 pandemic, with those living in vulnerable situations or from low socio-demographic backgrounds disproportionately affected. For example, young people are more likely to be affected by disruption in their education at a critical time and in the long-term most are at risk of poor employment and the associated health outcomes in economic downturn^[15]^. The psychosocial impacts of the COVID-19 pandemic on this population has been recognised, with charities and support groups offering online blogs, videos and support groups to mitigate the declines in mental health^[16]^. However, our findings highlight the importance of directing support to these groups during initial conversations with contact tracers. For example, as young people are more likely to be employed through the expanding gig economy, and have no access to sick pay, stable work contracts or the ‘furlough’ scheme^[24]^, it is important to consider support mechanisms so that these individuals do not fall through the cracks. A large multi-agency response is needed to deal with the wide range of needs identified in our study.

### The impact of self-isolation on women

Our findings suggest that women were nearly twice as likely as men to report mental health difficulties as a challenge to self-isolation. Currently, very little support exists that is targeted specifically at women during their self-isolation^[17]^. However, our study and others suggest that they evidence greater psychosocial harms than men. Around the world, the disproportionate impact of the COVID-19 pandemic on women has been well-recognised^[18]^. Women are over-represented in professions, especially nursing, working at the front-line of the response to COVID-19^[18]^, continue to do the majority of unpaid care within households^[19]^ and face high risks of economic insecurity^[18]^. Women also face an increased risk of violence, exploitation and abuse, particularly if they are living in a vulnerable situation during self-isolation^[20]^. Self-isolation and lockdown restrictions will also increase the demand for unpaid work at home, much of which traditionally falls onto women^[18]^.

Given that women are significantly more likely to face economic insecurity, live in vulnerable or harmful situations and be unpaid carers, support needs to be targeted specifically to this group. This support starts with raising awareness of the issue, by expanding community partnerships, training healthcare workers to recognise the signs of abuse and violence and spreading awareness about the importance of reporting incidents^[21]^. Internationally, to support these groups, countries have implemented warning systems to enable victims of gender and family violence to alert the authorities^[22]^. Recognising that, once again, a large multi-agency long-term response is needed to deal with the range of needs identified in our study.

### The impact of the COVID-19 pandemic on those on low incomes

Nearly two-thirds of our sample evidenced some degree of income precarity. People with high or very high income precarity were found to be more likely to report financial challenges, mental health concerns, loneliness, caring responsibilities, and a lack of support from family and friends, and work not supporting them to self-isolate as challenges to self-isolation, compared to those with low income precarity.

The COVID-19 pandemic has highlighted and exacerbated existing health inequalities, disproportionately impacting those who are in low-skilled employment, of younger age or have underlying health conditions^[23]^. Moreover, individuals who already struggled to cover their basic financial needs, were also significantly more likely to be placed on ‘Furlough’ during the pandemic^[24]^, further increasing their financial concerns and risk for non-compliance to COVID-19 restrictions. Importantly, these same groups who experience economic vulnerability are least likely to adhere to the behavioural and social interventions used to control the spread of COVID-19, such as social distancing and frequent hand washing ^[25-26]^. Evidence also highlights lower compliance with self-isolation guidance amongst these groups due to a wide range of contributing factors^[9, 25-26]^. Specifically, these low income groups (those who earn less than £20,000 a year or had less than £100 saved), report three times lower ability to self-isolate than their more wealthy counterparts^[26]^.

The financial impact of self-isolation has been recognised internationally, with Governments moving quickly to introduce schemes to compensate for financial loss during self-isolation to encourage adherence^[3]^. In Wales, the self-isolation support scheme was introduced on 23^rd^ October 2020^[27]^ and provided people who met eligibility criteria with £500 for each period of self-isolation up to a maximum of three times after they (or from 7^th^ December 2020 their child) had been advised to self-isolate^[28]^.

Identifying those with precarious incomes is important to halt the continuing spread of COVID-19 and mitigate its immediate economic and social harms. Given that our study found that people with precarious incomes not only experienced financial challenges of self-isolation, but also challenges with employers and mental health concerns, identification of people with precarious incomes is important to enable access to both financial and mental health support, as well as support to employers to enable employees to self-isolate. However, there are challenges associated with identifying these groups. Asking about income at the point of contact tracing could be viewed as sensitive, especially when efforts have been made by UK Governments to encourage engagement with the contact tracing systems by reassuring citizens that financial information, such as bank details, would not be requested. The use of an income precarity measure during initial conversations with contact tracers to direct financial and practical support could be an alternative and justified approach when set against the health and economic harms of self-isolation our study has observed with this group.

### Strengths and limitations

Our study is the first in Wales to examine the challenges faced by people asked to self-isolate. It included responses from a representative sample of contacts of COVID-19 identified through the national TTP database, ensuring that all those in the study had been confirmed and informed to self-isolate. This enabled sampling to ensure that contacts in our study were representative of all contacts successfully reached by TTP in each of the periods of interest for the two survey rounds. However, this also presents the key limitation of the study, as it includes only those who have been successfully reached by a contact tracer and informed to self-isolate. It is likely that those who are not successfully reached may perceive additional challenges to self-isolation that are not be captured in our findings.

## Conclusion

Our study found that several groups who are more likely to report facing challenges during periods of self-isolation to control the spread of COVID-19. These groups are specifically those with young adults, females and those with precarious. As vaccine coverage increases and societal restrictions on contacts are eased it is likely that there will be changes to the requirement for self-isolation. Yet, with the emergence of new variants of concern it remains an essential health protection measure for this and future pandemics. Identifying those with precarious incomes and at greatest risk of experiencing mental health harms of self-isolation during initial conversations with contact tracers is a key priority to ensure comprehensive assessment of needs and access to targeted support to help maximise potential adherence to self-isolation.

## Data Availability

The data that support the findings of this study are available on request from the corresponding author [KI]. The data are not publicly available due to privacy or ethical restrictions.

## Author contribution

Kate Isherwood, Ph.D. (conceptualisation; methodology; project management; writing – original draft; data analysis; writing – editing); Richard Kyle, Ph.D. (conceptualisation; funding acquisition; methodology; project oversight; supervision; data analysis; writing – editing); Benjamin Gray, Ph.D. (writing – editing; data analysis); Alisha Davies, Ph.D. (conceptualisation; funding acquisition; methodology; writing – editing; project oversight).

## Acknowledgements

The authors would like to thank colleagues in the Research and Evaluation Division, Public Health Wales, for their support in completing this work, particularly James Bailey and Poppy Escritt. The authors would also like to thank Beaufort Research Limited, who conducted data collection.

## Footnotes

1 There are two different types of contact tracing. Forward contact tracing is used to identify individuals who may have been infected by the index case. Backwards contact tracing is used to identify the setting or event or primary case who infected the index case (Endo et al., 2021). Due to capacity and nature of the pandemic, forward contact tracing has predominately been conducted with backwards contact tracing used in the context of outbreak control.

